# Oticon Zircon RIC in Adults Aged 65 and Over: A 19-Month Single-Centre Prospective Follow-Up

**DOI:** 10.64898/2026.11.26.26369074

**Authors:** Carla Abbou, Malik Hammadi

**Author notes:** Corresponding author: Carla Abbou —.

## Abstract

**Background:** Hearing aid benefit in older adults is usually reported from short follow-ups. We describe 19-month outcomes in a prospective cohort fitted at a manufacturer-run clinic.

**Methods:** 34 adults aged ≥65 with bilateral symmetric sensorineural hearing loss were fitted bilaterally with the Oticon Zircon RIC (NAL-NL2, REM-verified). Visits at 1, 3, 6, 12 and 19 months. Primary outcome: HHIE-25 change baseline to 6 months. Secondary outcomes: APHAB, IOI-HA, QuickSIN, data-logged use, unaided PTA4. Outcomes were assessed by an audiologist who fitted no participant; the analysis was performed by a biostatistician who is not an Otic employee.

**Results:** 26/34 (76.5%) completed 19 months. HHIE-25 improved by 21.4 points at 6 months (95% CI 17.1–25.7, n=31, p<0.001). Among completers, 19-month scores remained improved vs baseline (−18.1) but rose slightly from 6 months (+3.5). Mean use 9.0 h/day at 6 months and 8.1 h/day at 19 months. APHAB problem scores improved by 10.5 to 16.6 percentage points. IOI-HA stable (∼27–28). QuickSIN benefit 1.8 dB at 6 months. In tipping-point analyses, the improvement from baseline to 19 months remained statistically significant even with a large extra penalty applied to dropouts.

**Conclusion:** Benefit and use were largely maintained to 19 months with a modest decline after 6 months. Causal attribution is not possible. Interpret cautiously given single-centre, manufacturer-funded design and absence of a control group.

## 1. Introduction

Hearing aid use and benefit can change over time; long-term follow-up in routine care is reported less often than short-term outcomes [1,10]. The Hearing Handicap Inventory for the Elderly (HHIE-25) is widely used to measure self-reported hearing handicap. This report documents whether benefit and wear time were maintained over ∼19 months under protocol-defined conditions (verified fittings, scheduled visits, outcome assessment separated from fitting) at the Otic Clinical Hearing Centre. It is a prospective single-arm observational cohort study presented as a preprint. Our primary objective was to estimate the change in HHIE-25 from baseline to 6 months; secondary objectives were to describe the maintenance of benefit and device use to 19 months.

## 2. Methods

### 2.1 Design, roles and blinding

Prospective single-centre observational cohort of consecutive patients fitted 10 February–21 March 2025. Written informed consent to take part in the study and for the use of their anonymised data in research reporting and the publication of aggregated results was obtained from all participants. Fitting audiologist (Carla Abbou) performed fittings, REM and data-log reading. A second audiologist (outcome assessor) who fitted no participant performed audiometry, QuickSIN and scored questionnaires (completed by participant alone, sealed). Fitter blinded to outcome scores until data lock. A biostatistician who is not an Otic employee (fixed-fee contract) analysed the data under a pre-specified statistical analysis plan (version 2.0.5, 3 March 2025).

### 2.2 Participants

41 screened, 34 enrolled. Exclusions: air–bone gap ≥15 dB (n=3), interaural asymmetry >15 dB (n=2), MoCA <22 [2] (n=1), declined (n=1). Inclusion: age ≥65; bilateral symmetric SNHL, better-ear PTA4 26–70 dB HL; MoCA ≥22; able to attend visits.

### 2.3 Device and fitting

Oticon Zircon RIC 312, bilateral, NAL-NL2 [3], firmware v3.4.2 /FitSuite 12.1. Receivers 60 dB (n=29) or 85 dB (n=5). Couplings: open 10, vented tulip 12, closed 12. First-time users (n=24) started at 90% target → 100% at 1 month (22 at 1 m, 2 at 3 m); experienced users (n=10) fitted at 100%.

### 2.4 Real-ear verification

Audioscan Verifit 2 (v4.28.1), ISTS at 50/65/80 dB SPL, daily calibration. Probe tube ∼5 mm from tympanic membrane. Criterion: ±5 dB of NAL-NL2 target at 250–4000 Hz at 65 dB SPL. REM repeated at 1 month, 12 months and after gain changes >3 dB.

### 2.5 Schedule and measures

Visits: fitting, 1, 3, 6, 12, 19 months (windows ±2 weeks at 1, 3 and 6 months; ±3 weeks at 12 and 19 months). Measures: HHIE-25 [4,11], APHAB [5], IOI-HA [6], QuickSIN [7], data-logged h/day, unaided PTA4.

### 2.6 Statistical analysis

Primary: paired t-test baseline vs 6-month HHIE-25 (complete cases). Linear mixed model (random intercept, categorical time) using all available data (MAR). Completers-only trajectories. Sensitivity analyses: (1) exclusion of out-of-window visits; (2) tipping-point analysis of the baseline-to-19-month change, in which missing 19-month HHIE-25 values for dropouts were replaced by their last observed value plus a fixed penalty δ (0 to 30 points in steps of 5). No multiplicity correction; no formal power calculation.

## 3. Results

### 3.1 Participant flow

34 enrolled → 33 at 3 m (1 relocation) → 31 at 6 m (1 death unrelated, 1 withdrawal) → 29 at 12 m (1 death unrelated, 1 lost to follow-up) → 26 at 19 m (1 relocation, 1 hospitalisation, 1 withdrawal). Completion 76.5%. Dropouts older (80.1 vs 73.6 y) and higher baseline HHIE (52.5 vs 42.2). Two of 153 follow-up visits (1.3%) fell outside the protocol window. One of these was the 6-month visit of a participant seen 18 days outside the window; excluding it, the primary result was unchanged (n=30; mean change −21.40, SD 11.98; 95% CI −25.87 to −16.93). The other, at 12 months (+24 days), does not affect the primary endpoint. Follow-up among the 26 completers averaged 580 days (median 581.5, range 565 to 592; about 19.0 months). Across all 34 participants, follow-up to the last observed visit totalled 16,782 person-days (45.9 person-years; mean 494 days).

### 3.2 Baseline characteristics

### 3.3 Real-ear verification

Pre-adjustment MAD 65 dB SPL: 6.5 (2.7) dB. Post-adjustment: 2.8 (1.0) dB at 65 dB; 3.3 (1.1) at 50 dB; 3.9 (1.3) at 80 dB. 33/34 (97.1%) met ±5 dB criterion. REM re-checks at 1 and 12 months remained within tolerance.

### 3.4 HHIE-25 outcomes

All available: Baseline 44.6 (19.2, n=34); 3 m 27.9 (16.7, n=33); 6 m 22.9 (16.9, n=31); 12 m 22.9 (16.1, n=29); 19 m 24.1 (16.6, n=26).

Primary (complete-case n=31): change baseline→6 m = −21.4 (SD 11.8), 95% CI −25.7 to −17.1, p<0.001.

Completers (n=26): baseline→19 m change = −18.1 (SD 12.7), 95% CI −23.2 to −12.9. Means: 42.2 → 25.2 → 20.6 → 22.2 → 24.1. Change 6→19 m: +3.46 (6.82), 95% CI 0.70–6.22, p=0.016.

Mixed model (all available data): 6-month change −21.4 (95% CI −24.8 to −18.0), consistent with complete-case result.

**Figure 1.**
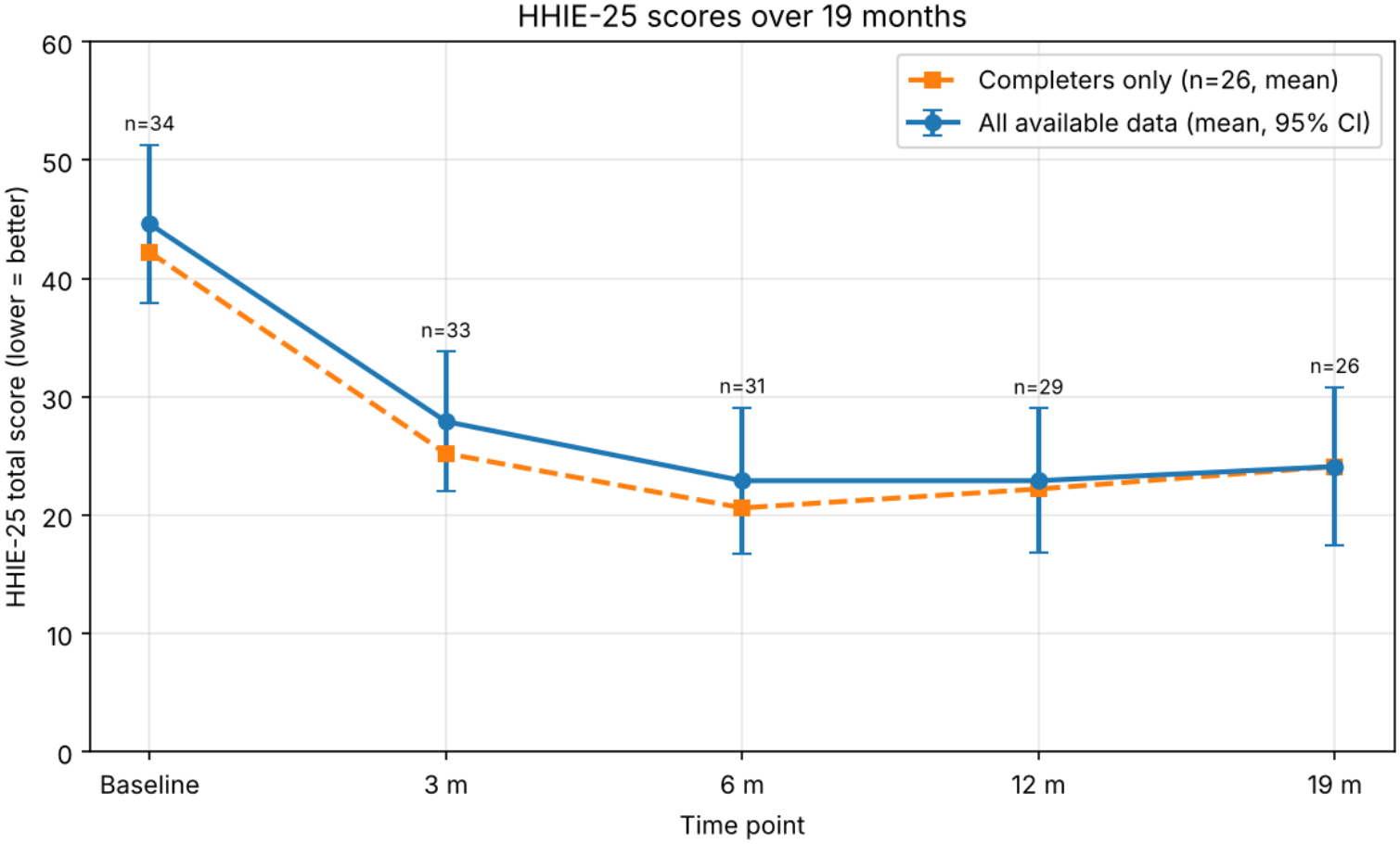
Mean HHIE-25 total score (lower = better) over 19 months. Blue: all available data with 95% confidence intervals; orange: the 26 completers.

### 3.5 APHAB

Baseline unaided (n=34): EC 51.1 (16.4), RV 47.8 (22.6), BN 60.6 (13.7). 6 m aided (n=31): EC 33.9, RV 37.1, BN 45.9. Paired benefit at 6 m: EC 16.6 (14.5), RV 10.5 (13.7), BN 13.6 (17.2). 12 m scores similar to 6 m.

### 3.6 IOI-HA

3 m 27.5 (4.4, n=33); 6 m 27.8 (3.9, n=31); 12 m 27.4 (4.2, n=29); 19 m 27.9 (3.7, n=26). Stable in the upper range throughout.

### 3.7 QuickSIN, use time and unaided audiometry

QuickSIN within-session benefit: 1.8 dB (95% CI 1.0–2.6) at 6 m; 1.7 dB (0.9–2.5) at 12 m. Mean use: 7.9 (1 m), 8.5 (3 m), 9.0 (6 m), 8.4 (12 m), 8.1 (19 m) h/day. Completers: −0.93 h/day (95% CI −1.81 to −0.06) from 6 to 19 m. Unaided better-ear PTA4 in completers worsened by 3.2 dB (95% CI 1.4 to 5.0) from baseline to 19 months.

### 3.8 Sensitivity analyses for missing data

Tipping-point analysis (n=34): missing 19-month HHIE-25 values for the 8 participants lost to follow-up were replaced by their last observed value plus a fixed penalty δ, and the baseline-to-19-month change for the full cohort was recalculated (Table 2). With a penalty of 30 points applied to every dropout, the mean improvement from baseline remained statistically significant (95% CI −17.20 to −4.33). Completers-only and mixed-model results were consistent with the primary analysis.

**Table 1.** Baseline characteristics. Severity categories by better-ear PTA4: mild 26–40, moderate 41–55 and moderately severe 56–70 dB HL.

| **Characteristic** | **n = 34** |
|---|---|
| Age, years, mean (SD) [range] | 75.1 (6.2) [65–88] |
| Female sex, n (%) | 18 (52.9%) |
| Better-ear PTA4, dB HL, mean (SD) | 43.8 (9.1) |
| Worse-ear PTA4, dB HL, mean (SD) | 47.6 (9.4) |
| Severity (better-ear PTA4) | Mild 12 (35.3%); Moderate 17 (50.0%); Mod. severe 5 (14.7%) |
| First-time / Experienced users | 24 (70.6%) / 10 (29.4%) |
| MoCA, mean (SD) | 25.7 (1.7) |
| Baseline HHIE-25, mean (SD) | 44.6 (19.2) |
| Coupling (open / vented tulip / closed) | 10 / 12 / 12 |

**Table 2.** Tipping-point analysis of the baseline-to-19-month HHIE-25 change (n=34; missing 19-month values replaced by last observation plus δ).

| **δ (extra points)** | **Mean change, baseline→19 m** | **95% CI** | **Benefit maintained?** |
|---|---|---|---|
| 0 | −17.82 | −22.25 to −13.40 | Yes |
| 5 | −16.65 | −21.17 to −12.13 | Yes |
| 10 | −15.47 | −20.20 to −10.74 | Yes |
| 15 | −14.29 | −19.34 to −9.25 | Yes |
| 20 | −13.12 | −18.56 to −7.68 | Yes |
| 25 | −11.94 | −17.85 to −6.03 | Yes |
| 30 | −10.76 | −17.20 to −4.33 | Yes |

## 4. Discussion

Substantial HHIE-25 improvement by 6 months was largely maintained to 19 months, with a small rise in scores thereafter that was nominally significant in this exploratory comparison. The later rise of 3.5 points was small compared with the initial improvement of 21.4 points. In tipping-point analyses the improvement from baseline to 19 months remained statistically significant even with a 30-point extra penalty applied to dropouts (Table 2). Daily use remained high. Findings are in line with prior longer-term observational reports [1,10]; this study adds REM-verified fittings and outcome assessment separated from fitting, in a manufacturer-run setting. The aided QuickSIN benefit of 1.8 dB was modest. Unaided better-ear PTA4 worsened by 3.2 dB over 19 months, of a similar order to age-related change in longitudinal studies [9]; measurement error should be considered when interpreting small threshold changes [8]. The uncontrolled design precludes causal attribution: natural fluctuation, ageing, regression to the mean and expectation effects may all contribute. Results are best viewed as descriptive real-world evidence.

## 5. Limitations

- No control group — causal inference impossible.
- Open-label, manufacturer-funded, single-centre; the study author and outcome assessor are employed by the manufacturer. Outcome assessment separated from fitting, but residual bias possible.
- Attrition 23.5% non-random. Tipping-point analyses support robustness of the improvement from baseline to 19 months; residual selection bias remains possible.
- Small sample, no formal power calculation, no multiplicity correction.
- MoCA <22 excluded; single-device setting limits generalisability.
- Data logging = device-on time, not listening time.
- Preprint — not peer-reviewed.

## 6. Conclusion

Self-reported handicap improved substantially by 6 months and remained improved at 19 months among completers with high daily use, despite a modest decline after 6 months. Findings are compatible with maintained improvement under protocol-driven, REM-verified fitting, although this study cannot show that the protocol or the verification caused it. The improvement from baseline to 19 months remained statistically significant in tipping-point sensitivity analyses. Interpretation is limited by lack of control group, attrition, sample size and manufacturer involvement. These are descriptive real-world observations.

## Data Availability

All data produced in the present study are available upon reasonable request to the authors

## Declarations

### Ethics and governance

The study was reviewed by the Comité de Protection des Personnes (CPP) Sud-Ouest et Outre-Mer, which gave a favourable opinion on 23 January 2025 (reference 435655; ID-RCB 2025-A00763-49), before enrolment began on 10 February 2025. All participants provided written informed consent to take part in the study and for the use of their anonymised data in research reporting and the publication of aggregated results. Data were pseudonymised during the study and anonymised after data lock, and access was restricted to the study team. Processing complied with the GDPR (EU 2016/679) and the Loi Informatique et Libertés, under the supervision of the Commission Nationale de l’Informatique et des Libertés (CNIL).

### Trial registration

The study was registered on ClinicalTrials.gov (NCT06860672) on 13 March 2025, after enrolment began on 10 February 2025 (retrospective registration).

### Data availability

Individual participant data are not publicly available. Participants consented to the publication of aggregated results only, and individual records from a small single-centre cohort of older adults could allow re-identification. Aggregated summary data supporting the findings are reported in the article. Requests for further aggregated data may be sent to the corresponding author and will be considered subject to review by the data protection officer of Otic and a data-sharing agreement.

### Author contributions

Carla Abbou: conception, data collection, fitting, analysis oversight, drafting. Dr Malik Hammadi: senior supervision, critical revision, final approval. Both approved the preprint.

## Acknowledgements

The authors thank Jean-Pierre Moreau (outcome assessor) and François Laurent (biostatistician, ISPED, University of Bordeaux).

## Generative AI

Generative AI tools, including Claude (Anthropic), were used to assist with literature checks, statistical verification, review of the manuscript against medRxiv requirements, manuscript structuring, drafting and language editing, figure generation and preparation of the STROBE checklist. No generative AI system performed the primary statistical analysis or generated original study results. The authors reviewed all content and take full responsibility for it.

## STROBE statement

This report follows the STROBE guidelines for observational studies. A completed STROBE checklist is provided in the Appendix.

## APPENDIX STROBE Checklist (Cohort Study)

STROBE Statement — checklist of items that should be included in reports of observational studies (Cohort version). Completed for the present manuscript.

| **Item No.** | **Recommendation** | **Reported?** | **Location in manuscript** |
|---|---|---|---|
| 1a | Indicate study design in title or abstract | Yes | Title + Abstract |
| 1b | Informative, balanced abstract | Yes | Abstract |
| 2 | Background / rationale | Yes | Section 1 |
| 3 | Objectives / hypotheses | Yes | Section 1 |
| 4 | Key elements of study design | Yes | Section 2.1 |
| 5 | Setting, locations, dates | Yes | Sections 2.1–2.2 |
| 6a | Eligibility criteria, sources and methods of selection; methods of follow-up | Yes | Sections 2.2, 2.5, 3.1 |
| 6b | Matching criteria (matched studies only) | N/A | No matching performed |
| 7 | Define outcomes, exposures, predictors, confounders | Yes | Sections 2.5–2.6 |
| 8 | Data sources / measurement | Yes | Sections 2.4–2.5 |
| 9 | Efforts to address bias | Yes | Sections 2.1, 2.6, 5 |
| 10 | Study size explanation | Yes | Sections 2.2, 2.6 (34 consecutive participants; no formal power calculation) |
| 11 | Quantitative variables handling | Yes | Sections 2.5–2.6 |
| 12a | Statistical methods (incl. confounding) | Yes | Section 2.6 |
| 12b | Subgroups / interactions | N/A | No subgroup or interaction analyses performed |
| 12c | Missing data methods | Yes | Sections 2.6, 3.8 |
| 12d | Loss to follow-up (cohort) | Yes | Sections 3.1, 3.8 |
| 12e | Sensitivity analyses | Yes | Section 3.8 |
| 13a | Numbers at each stage | Yes | Section 3.1 (flow) |
| 13b | Reasons for non-participation | Yes | Section 3.1 |
| 13c | Flow diagram considered | Partly | Flow described in text (Section 3.1); no separate diagram |
| 14a | Characteristics of participants | Yes | Table 1, Section 3.2 |
| 14b | Indicate number with missing data | Yes | Sections 3.1, 3.4–3.7 (n at each time point) |
| 14c | Follow-up time (cohort) | Yes | Section 3.1 |
| 15 | Outcome data | Yes | Sections 3.4–3.7 |
| 16a | Unadjusted / adjusted estimates + precision | Yes | Sections 3.4–3.8 (unadjusted estimates with 95% CIs; no confounder adjustment) |
| 16b | Category boundaries reported | Yes | Table 1 caption |
| 16c | Relative + absolute risks if relevant | N/A | Not applicable |
| 17 | Other analyses (subgroups, sensitivity) | Yes | Section 3.8 |
| 18 | Key results summarised | Yes | Abstract, Section 4 and Section 6 |
| 19 | Limitations (bias, imprecision) | Yes | Section 5 |
| 20 | Overall interpretation | Yes | Sections 4 & 6 |
| 21 | Generalisability | Yes | Sections 4 & 5 |
| 22 | Funding sources | Yes | Declarations |

*Note: This completed checklist follows the official STROBE recommendations for cohort studies (von Elm et al., PLoS Medicine 2007; www.strobe-statement.org). Page/section references correspond to the present manuscript.*

## References

[1] Hickson L, et al. Factors associated with success with hearing aids in older adults. Int J Audiol 2014;53(Suppl 1):S18–27.

[2] Nasreddine ZS, et al. The Montreal Cognitive Assessment, MoCA. J Am Geriatr Soc 2005;53:695–9.

[3] Keidser G, et al. The NAL-NL2 prescription procedure. Audiol Res 2011;1:e24.

[4] Ventry IM, Weinstein BE. The Hearing Handicap Inventory for the Elderly. Ear Hear 1982;3:128–34.

[5] Cox RM, Alexander GC. The Abbreviated Profile of Hearing Aid Benefit. Ear Hear 1995;16:176–86.

[6] Cox RM, et al. Norms for the International Outcome Inventory for Hearing Aids. J Am Acad Audiol 2003;14:403–13.

[7] Killion MC, et al. Development of a quick speech-in-noise test. J Acoust Soc Am 2004;116:2395–405.

[8] Schmuziger N, et al. Test-retest reliability of pure-tone thresholds. Ear Hear 2004;25:127–32.

[9] Lee FS, et al. Longitudinal study of pure-tone thresholds in older persons. Ear Hear 2005;26:1–11.

[10] Takahashi G, et al. Subjective measures of hearing aid benefit and satisfaction in the NIDCD/VA follow-up study. J Am Acad Audiol 2007;18:323–349.

[11] Weinstein BE, Spitzer JB, Ventry IM. Test-retest reliability of the Hearing Handicap Inventory for the Elderly. Ear Hear 1986;7:295–299.

